# Detection of circulating plasma tumour mutations in early stage triple negative breast cancer as an adjunct to pathological complete response assessment

**DOI:** 10.1101/2023.05.31.23290797

**Authors:** Elena Zaikova, Brian Y.C Cheng, Viviana Cerda, Esther Kong, Daniel Lai, Amy Lum, Cherie Bates, Wendie den Brok, Takako Kono, Sylvie Bourque, Angela Chan, Xioalan Feng, David Fenton, Anagha Gurjal, Nathalie Levasseur, Caroline Lohrisch, Sarah Roberts, Tamara Shenkier, Christine Simmons, Sara Taylor, Diego Villa, Ruth Miller, Rosalia Aguirre-Hernandez, Samuel Aparicio, Karen Gelmon

**Affiliations:** Molecular Oncology, BC Cancer, 675 W10th Avenue, Vancouver; Medical Oncology, BC Cancer, 600 W10th Avenue, Vancouver; Medical Oncology, BC Cancer, 13750 96 Ave, Surrey; Medical Oncology, BC Cancer, 2410 Lee Ave, Victoria; Medical Oncology, BC Cancer, 32900 Marshall Rd, Abbotsford; Medical Oncology, BC Cancer, 1215 Lethbridge St, Prince George; Medical Oncology, BC Cancer, 399 Royal Ave, Kelowna; Imagia Canexia Health, 204-2389 Health Sciences Mall, Vancouver

## Abstract

Circulating tumour DNA (ctDNA) detection in liquid biopsy is an emerging alternative to tissue biopsy, but its utility in treatment response monitoring and prognosis in triple negative breast cancer (TNBC) is not yet well understood. In this study, we determined the presence of ctDNA detectable actionable mutations with a clinically validated hotspot treatment indication panel in early stage TNBC patients, without local recurrence or metastatic disease at the time of evaluation. Sequencing of plasma DNA and validation of variants from 130 TNBC patients collected within 7 months of primary treatment completion revealed that 7.7% had detectable residual disease with a hotspot panel. Among neoadjuvant treated patients, we observed a trend where patients with incomplete pathologic response and positive ctDNA within 7 months of treatment completion were at much higher risk of reduced progression free survival. We propose that a high risk subset of early TNBC patients treated in NAT protocols may be identifiable by combining tissue response and sensitive ctDNA detection.

## Introduction

Breast cancer is the most commonly diagnosed cancer worldwide and accounts for nearly 25 % of newly diagnosed cancers in women^1^. Triple negative breast cancers (TNBC) account for approximately 15 % of new breast cancer diagnoses and are associated with poorer prognosis and earlier disease progression than other cancer types^2,3^. The ability to monitor disease progression as well as response to treatment in TNBC patients would provide physicians with an invaluable tool to help tailor treatment to the individual and improve patient outcomes^4^. Detection of circulating tumour DNA (ctDNA) is an emerging non-invasive alternative to tissue biopsy approaches, as it only requires a blood sample and can therefore be collected at different timepoints with minimal discomfort to the patient. ctDNA can be found in the blood of patients with all stages of disease and carries many of the features of the solid tumour^5–7^. Consequently, ctDNA can be used in screening and early detection, disease monitoring, recurrence prediction, tumour profiling, and informing treatment sequence in many solid cancers^8–13^. ctDNA surveillance has the benefit of detecting recurring disease months before imaging findings are present^14,15^. With current surveillance recommendations after treatment completion not routinely imaging asymptomatic patients for distant recurrence, routine ctDNA monitoring may have a high probability of capturing participants with aggressive disease progression. This could lead to earlier treatment of minimal residual disease and studies to assess the utility of this strategy. ctDNA detection approaches vary between studies and often rely on personalised mutational profiling, presenting challenges in assay scalability and generalizability of detection methodologies. A prognostic signal for ctDNA in TNBC patients has been suggested from earlier studies^9,16–18^, however the detection rate and outcome associations vary depending on the implementation of the specific research grade or clinical test and the clinical stage of the patients. Using a clinically validated targeted mutation hotspot ctDNA sequencing panel, designed for actionable therapy guidance, here applied to early stage TNBC including clinically disease free patients, we report that ctDNA evaluation in a 7 month interval after completion of primary treatment may identify a subgroup of TNBC patients who are at high risk of relapse.

## Materials and methods

### Clinical study design and participant recruitment program

Materials used for this study were obtained under the Precision Medicine for Breast Cancer Research program, a British Columbia-wide breast cancer participant recruitment program focused on collecting high-quality human samples for genomic and translational research. Eligible TNBC participants with early stage primary TNBC were recruited into the TNBC exploratory study (REB approval H15-01764). At the time of consent, a baseline blood draw and saliva sample was obtained alongside FFPE tissue materials from primary tumour surgical resection or core biopsy, followed by subsequent blood draws between completion of primary adjuvant or neoadjuvant treatment protocols. Consent for use of surgical and core biopsies and clinical records data was obtained. All participants provided informed consent, and the study design followed good clinical research practices outlined by the Office of Biobank Education and Research (OBER). Special care was taken to ensure participant data and all corresponding clinical information were coded to protect confidentiality, and ctDNA results were either anonymized or withheld from collaborating oncologists, pathologists and researchers.

### Participant sample and clinical data collection

Research staff trained on the protocol at each participating BC Cancer centre identified eligible TNBC participants and obtained informed consent. Participants were 18 years or older, with a diagnosis of TNBC. Pregnant participants and those with a history of previous invasive cancer of DCIS were included. Sufficient primary tumour tissue, fresh or FFPE, was available for all included participants. Exclusion criteria included known blood disorders or a history of other solid or hematologic malignancy within the preceding 5 years, except for appropriately treated CIS of the cervix, Stage I uterine cancer and non-melanoma skin carcinoma. The research study was explained to the participants in a private medical setting and sufficient time was given to explain the protocol and have their questions answered.

For enrolled participants, treatment naive TNBC tumour samples were requested from hospitals for pathology review and DNA extraction. The source of tumour samples were either fresh frozen tumour tissue, diagnostic core biopsy blocks, or chemo-naive surgery blocks, and corresponding H&E slides were reviewed by a pathologist to identify invasive tumour for DNA extraction. Core biopsy samples were sectioned at 10 µm thickness, while surgery samples had one to two 1-mm cores punched in pathologist-circled regions.

Saliva samples were collected with assistance and in-person instructions from dedicated clinical research staff. Oragene saliva collection kits were used to stabilize the sample before DNA extraction. Participant peripheral blood was collected in Streck Cell-Free DNA Blood Collection tubes (BCTs), and processed by a dedicated, trained biobank technician. In order to ensure high quality cfDNA samples free of lymphocyte-derived genomic DNA, Streck BCTs were chosen as the blood collection tube for their validated lymphocyte stabilizing solution^19^. Participant blood samples were then accessioned, anonymised, and processed within 10 days post-phlebotomy as previously described^19^ with further modifications to isolate buffy coat cell pellets.

### DNA extraction and sequencing

DNA extraction was done using commercially available DNA extraction kits in accordance with manufacturer protocols. For saliva DNA, the entire volume in the Oragene saliva collection tube was used for DNA extraction with the Oragene saliva extraction kit, eluting saliva DNA in ∼200 µL of TE buffer. To separate whole blood into its constituent layers, samples were centrifuged in a free-bucket centrifuge at 1,600 x g for 15 minutes. The top plasma layer was aspirated and aliquoted for long-term storage in vapour phase nitrogen tanks. The buffy coat layer was collected and further purified through erythrocyte lysis and phosphate-buffered saline washes, and resuspended in cell freezing media (50% RPM1, 40 % FBS, 1 0% DMSO) for storage at -80 °C.

Cell-free DNA was extracted from 2-4 mL of plasma using either the column vacuum-based QIAGEN Circulating Nucleic Acid kit or the magnetic bead-based cfDNA extraction kit, eluting DNA in 50-60 µL of AVE buffer or dH^2^0 respectively. Tumour FFPE DNA was extracted with the QIAGEN FFPE Extraction Kit, using the recommended QIAGEN deparaffinization solution and eluting DNA in 30 µL of TAE buffer. All extracted DNA samples were quantified with the Qubit dsDNA assay (ThermoFisher Scientific), and temporarily stored at -20 °C prior to sequencing and at -80 °C for long term storage.

Although a large majority of plasma DNA samples showed no genomic DNA contamination, routine QC was performed to ensure cfDNA sample quality. Using the Agilent 2100 Bioanalyzer and Agilent DNA High Sensitivity Kit, cfDNA with genomic contamination were identified by distinct high bp fragments (> 10,000 bp) in the electropherogram. Only samples with no genomic DNA contamination continued to amplicon panel sequencing.

Targeted sequencing was performed using the commercially-available Follow-It® amplicon panel (Imagia Canexia Health, previously known as Contextual Genomics Inc.) and the Find-It® target amplicon panel previously described^20^. The Follow-It® and Find-It® amplicon sequencing assays target a set of pan-cancer, clinically actionable hotspot SNV and small indel (up to 24 bp) mutations specifically designed for ctDNA and FFPE tumour DNA. Several modifications to the Find-It® panel were made by Imagia Canexia Health, including the addition of three target genes: MAP2K1, EGFR and POLE, and the removal of five target genes: STK11, PTEN, JAK1, FGFR1 and FGFR2 (https://imagiacanexiahealth.com/solution/plasma-follow-it/).

Briefly, post-treatment plasma and matched buffy coat DNA samples from 130 participants, along with matched tumour and saliva DNA for a subset of 60 participants (Figure 1A), were amplified using the Follow-It® PCR primer panel, generating an intermediate PCR product, which was subsequently purified using Ampure XP beads. The purified intermediate PCR product was then indexed with Illumina XT v2 adapters. Indexed PCR products were size-selected with Ampure XP beads to produce a final cleaned-up library of ∼275 bp fragments. Multiplexed libraries were pooled in equimolar concentrations and sequenced on the Illumina MiSeq platform using the V2 300 cycle kit, to a mean depth of coverage of ∼7500X.

**Figure 1.**
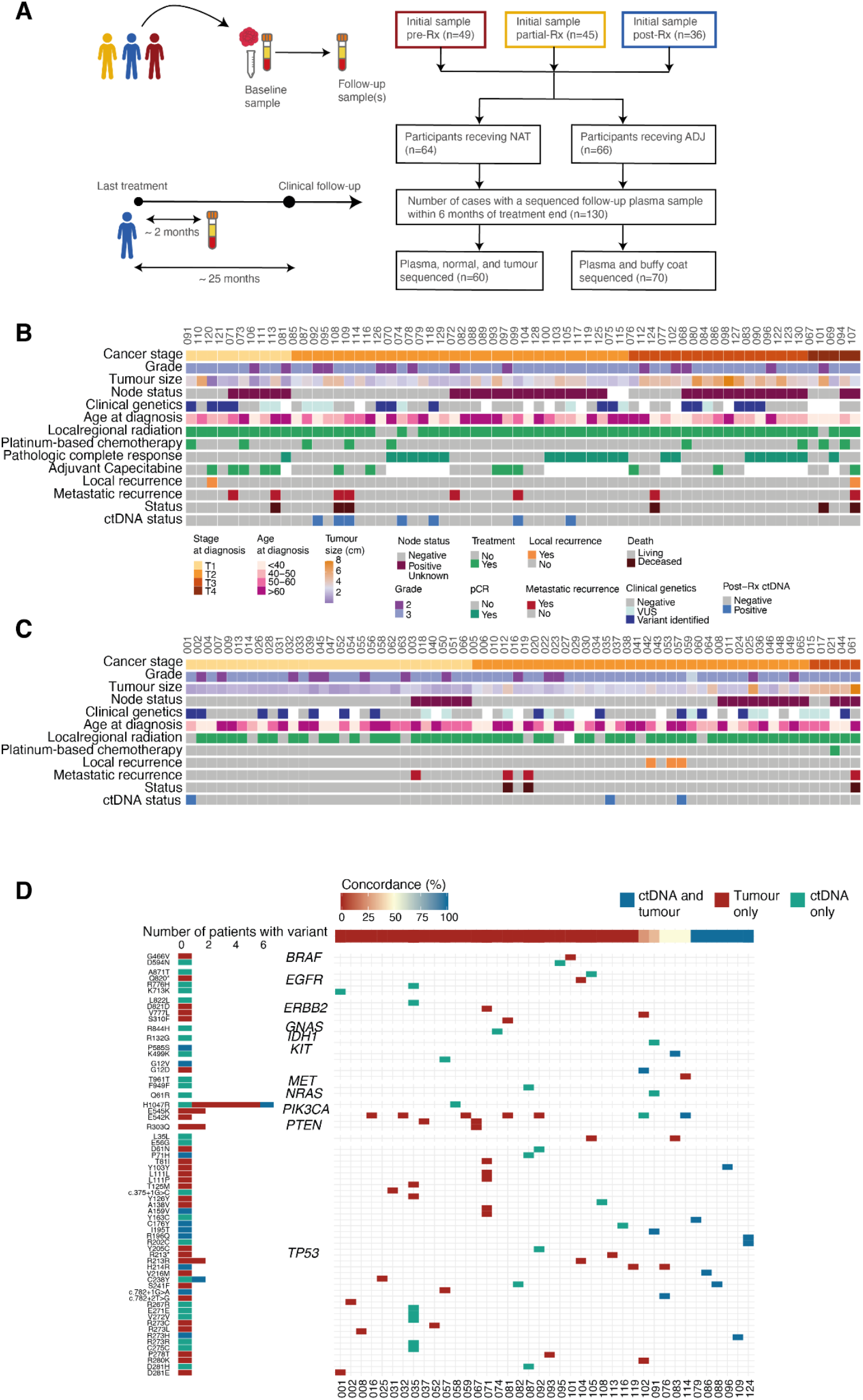
Sample collection and cohort overview. (**A**) Schematic of sample collection and sequencing summary for participants with a post-treatment sample collected within 7 months of treatment completion. (**B**) Clinical characteristics of NAT participants. (**C**) Clinical characteristics of ADJ participants. (**D**) Participant-specific concordance between somatic variants identified in FFPE tumour and plasma samples for participants with a detectable panel-targeted mutation.

### Identification of variants in target regions

Proprietary quality assurance methods based on DNA sequence barcodes that were incorporated into the assay and the bioinformatics pipeline, were used to increase sensitivity of called mutations. The bioinformatics pipeline first removed poor quality reads based on sequence length and base mismatches in the primer region. Reads passing the quality control filters were then aligned to the hg19 reference genome. Mutations were called with a supervised classification method that returned the random forest (RF) probability score of a variant belonging to the mutation class as opposed to the artifact class, with classification based on the alignment, sequence composition, and barcode information of the variant. Identified variants were then annotated using SnpEff v 4.3^21^.

Variant allele frequency (VAF) of identified variants was calculated using TA/(TA+TR+TO) where TA is the number of reads with second alternative allele, TR is the number of reads with reference allele, and TO is number of reads with “other” alleles, that is, third and fourth alternative alleles, and presented as a percentage.

### Validation of low-abundance variants by ddPCR

Samples with hotspot mutations with VAF below 1.5 % and above 0.5 % found through the Follow-It® sequencing panel were then independently validated using a Droplet Digital PCR (ddPCR) system for internal QC and bioinformatics purposes, where validated assays were commercially available. Additionally, variants with high VAF were validated with ddPCR to confirm detection of high-confidence variant calls (Figure S1A). In brief, lab-validated genotyping probe assays were purchased from BioRad and used to detect low-prevalence SNV mutations in ctDNA. In accordance with standard ddPCR workflow, PCR reactions were partitioned into picoliter sized droplets using the QX200 Droplet Generator (Bio-Rad Laboratories), and amplified using the manufacturer recommended number of PCR cycles for each assay. End-point droplet quantification was then performed using the QX200 Droplet Reader (Bio-Rad Laboratories), with mutation positive samples requiring at least 3 separate FAM positive only droplets and fluorescence signals 10-times higher than the associated wild-type only control. Positive wild-type and positive mutant control were amplified in conjunction with our ctDNA sample, using either cell line DNA, Horizon HD780 multiplex ctDNA reference standard or gene-blocks as positive controls (Table S1).

### Identification of participant-specific ctDNA signatures and statistical analyses

Single nucleotide variants and short indels identified by analysis of target panel sequencing and subsequent classification were further filtered prior to downstream analyses. First, variants with a RF probability score < 0.7 were removed, as were those with VAF below 1 %, based on ddPCR results. Likely germline variants and those resulting from clonal hematopoiesis were then identified and excluded from further analysis. This was done by comparing variants in saliva and buffy coat with variants in plasma and primary tumour samples in a participant-specific manner. Next, variants outside of the panel target regions were excluded from analyses. Finally, variants confirmed by ddPCR were added back into the pool of detected variants, whereas those with negative ddPCR results were excluded from downstream analyses. Additional filtering of variants with VAF > 40 % was performed to remove likely germline variants.

To compare clinicopathological characteristics between participants receiving neoadjuvant chemotherapy and participants treated with adjuvant chemotherapy, chi-square and Wilcoxon tests were used for categorical and continuous variables, respectively. Kaplan-Meier survival probability, faceted by whether ctDNA was detected within 7 months post-treatment was calculated using the survminer R package. Data presentation and analyses were performed using R version 3.6.1 and visualized using ggplot2, UpSetR, survminer, ggvenn, cowplot and patchwork R packages^22,23^.

## Results

### Cohort description

We measured ctDNA mutations detected with a clinically actionable ctDNA panel in plasma collected within 7 months following the completion of primary treatment in a cohort of 130 participants with non-metastatic TNBC, with no clinical progression at time of sample collection. The cohort comprised 64 participants who received neoadjuvant chemotherapy (NAT) followed by surgery, and 66 participants who received surgery followed by adjuvant chemotherapy (ADJ) (Figure 1B and C). The median age of participants at diagnosis was 54 years, and was similar across NAT and ADJ participants (Figure 1 and Table S2). Similarly, tumour grade and rate of variant detection by clinical genetics were similar across participants receiving NAT and ADJ treatment. Variants detected by clinical genetics included BRCA mutations in 8 ADJ and 13 NAT participants. The median follow-up interval between the date of last treatment and the sequenced post-treatment samples was 1.8 months with no significant difference between NAT and ADJ participants (Table S2). Following treatment completion, the median clinical follow-up was 25 months, with a range of 1 to 53 months. Similarly, rates of disease progression or death and intervals between treatment completion and recurrence, as well as time between treatment completion and disease progression or death were not statistically different between NAT and ADJ treated participants, despite overall sooner progression in participants receiving NAT (Table S2). Lymphovascular invasion was present in a higher proportion–21%–of ADJ participants than NAT (9%). There was a statistically significant difference in lymph node status between NAT and ADJ participants; 55% NAT participants had positive node status, compared to 27% of ADJ participants (Figure 1 and Table S2). Although half of all NAT and ADJ participants had stage T2 tumours at time of presentation, NAT participants tended to have a higher tumour stage (34% of NAT participants with tumour stage T3 or T4) than ADJ participants, 42% of whom had a T1 tumour stage. Similarly, ADJ participants tended to have smaller tumour sizes than participants receiving NAT (Figure 1, Table S2). Chemotherapy protocols differed between NAT and ADJ participants, with 9 (14%) NAT participants receiving chemotherapy containing platinum, while only 1 ADJ participant received platinum-containing therapy. Additionally, a higher number of ADJ participants received dual therapy compared to NAT. Most (87%) of all the participants in the cohort received local regional radiation; however, fewer NAT participants did not receive this treatment compared to ADJ (4 and 12 participants, respectively). Pathological complete response (pCR) following neoadjuvant therapy was achieved in 24 (38%) of NAT participants. Of the 40 NAT participants without pCR, 13 received additional adjuvant Capecitabine after surgery (Figure 1B). In addition to chemotherapy, surgery and radiation, 7 ADJ and 9 NAT participants received hormone/ zoledronic acid treatments.

To investigate whether we could detect residual disease following treatment completion, we used a sensitive clinically-validated ctDNA PCR sequencing panel^20,24^ and an accompanying mutation calling algorithm, covering 337 hotspots in 38 genes, to detect actionable mutations in ctDNA samples collected within 7 months of primary disease treatment completion from all 130 participants. To help define criteria for variant filtering and inclusion, we additionally sequenced matched buffy coat samples for all participants, as well as ctDNA from earlier and later time points, FFPE and saliva samples for subsets of the cohort.

First, we established the variant allele frequency (VAF) range for specific mutation detection by confirming 48 detected ctDNA variants with a probability score > 0.7 (random forest determined detection probability^20^) that had validated, commercially available (Biorad) digital droplet (ddPCR) assays. These 48 variants ranged from 0.1 % to 78.4 % VAF and were distributed across 8 genes and encompassed 24 different mutations (Table S1). A total of 14 variants were confirmed, while 34 were not detected by ddPCR (Figure S1A). The confirmed variants (true positives) were in 3 genes, PIK3CA, TP53 and KRAS, and had a median panel-estimated VAF of 1.1 % (VAF range: 0.5 % - 78.4 %). The median VAF for false positives was 0.7 % (VAF range: 0.1 % - 19.4 %) (Figure S1A). These results were used to establish for 1 % VAF or above criteria for variant filtering subsequent analyses. All true positives were included in downstream analyses, and all false positives were excluded.

### Detection of actionable ctDNA variants

Variants meeting the filtering criteria were distributed across 14 genes, with POLE and MAP2K2 variants detected in normal samples only, representing likely germline variants (Figure S1B). We compared buffy coat and ctDNA variants for all 130 participants to identify and exclude germline variants. Just 5 variants were detected in buffy coat samples, only 1 (TP53 R248G, VAF ∼1 %) of which was concordant with a participant-specific plasma sample (Figure S1C). Clonal hematopoiesis (CH) can contribute to false positives in ctDNA genotyping^25^. Therefore, we compared variants from saliva, lymphocyte, plasma, and FFPE tumour DNA from a subset of 60 participants to identify potential CH. We defined a CH mutation as any variant detected in plasma that is concordant with buffy coat but not with saliva. Out of the 93 variants detected in this subset of 60 patients, none were CH mutations as defined above (Figure S1C). Indeed, there was only 1 variant detected in a buffy coat sample within this cohort subset–the likely synonymous benign germline mutation EGFR R836R (rs2229066) (Figure S1C).

After excluding likely germline variants, we examined the concordance of detected variants in matched FFPE tumour and plasma samples for 60 participants. A total 72 variants were detected in tumour and plasma: 36 were detected in tumour samples only, 24 in plasma only, and 12 variants were present in both. Of the 60 participants with matched tumour and plasma sequences, 41 had a detectable variant in at least one source, 11 of which had a detectable variant in both sources (Figure 1D). Of these, 6 participants had 100% tumour-plasma concordance, with all detected variants in TP53 hotspots (Figure 1D). Of participants with a variant in plasma and/or tumour, 24 had a mutation in TP53, including 9 of the 12 concordant variants, 71% of the plasma only variants, and 51% of the tumour only variants (Figure 1D and Figure S1B). The next most common gene with tumour variants was PIK3CA (Figure S1B). Among the 12 concordant variants, VAF was higher in tumour samples than ctDNA, with a median VAF of 32.9% and 1.6% for tumour and plasma variants, respectively (Figure S1D). The most prevalent solid tumour mutation was PIK3CA H1047R, which was detected in tumour samples from 6 participants and was 1 of just 2 plasma variants found in more than 1 participant (each was detected in 2 participants) (Figure 1D). This pathogenic missense mutation has previously been reported to be associated with lower pathological complete response in TNBC participants treated with NAT^26,27^. The distribution of plasma and tumour variants in our cohort reflects previously reported population frequencies of somatic SNVs in TNBC^28–34^.

### Composition of post-treatment ctDNA variants

Actionable hotspot mutations were detected in post-treatment 7 month follow-up plasma in 10 of the 130 (7.7%) participants, which included 7 NAT and 3 ADJ (Figures 1B and C). Four participants with post-treatment ctDNA were subsequently diagnosed with recurrence. The distribution of detected mutations in post-treatment samples showed 13 variants in 6 genes: 6 TP53 variants from 4 participants, 3 EGFR variants from 3 participants, and 1 variant in each of ERB2, GNAS, KIT, and PIK3CA (Figure 2A). Two of the 6 TP53 variants detected in participant ctDNA following treatment were intron variants predicted to have high functional impact and three were missense variants predicted to have moderate functional impact (Figure 2A). The median VAF for TP53 variants of 2.76 % was higher than that of mutations in GNAS (2.1 % VAF), EGFR (median 1.4 % VAF), ERB2 (1.0 % VAF), KIT (1.3 % VAF) and PIK3CA (0.6 % VAF) variants. Additionally, the TP53 intron variants predicted to have high functional impact had VAFs exceeding 10% (Figure 2A). Three of the four participants with recurrence and post-treatment ctDNA had TP53 variants.

**Figure 2.**
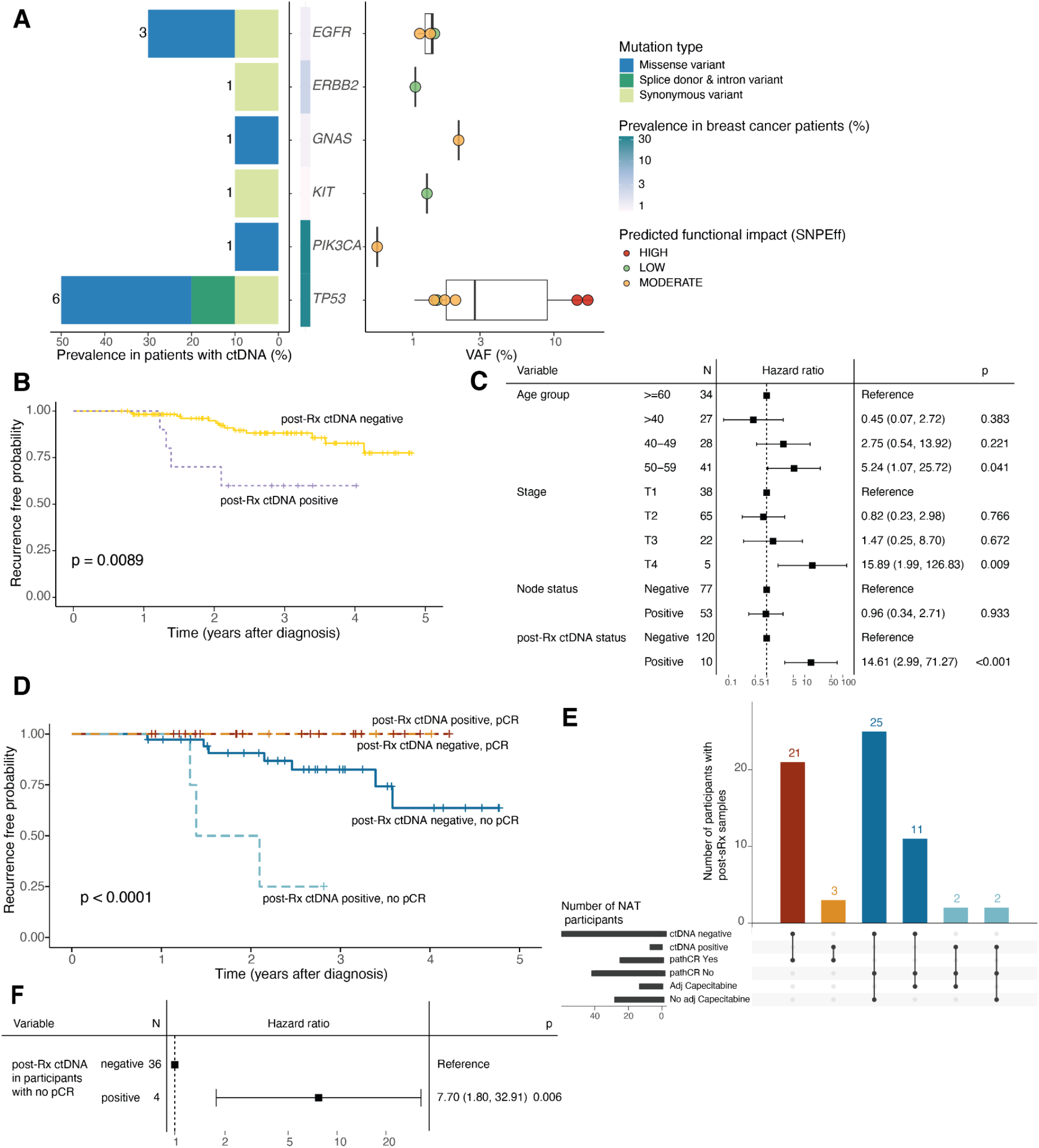
Recurrence-free survival and ctDNA in post-treatment plasma collected within 7 months of primary treatment completion in TNBC participants. (**A**) Distribution of detected somatic ctDNA variants across target genes. (Left) Prevalence of somatic mutations in patients with post-treatment ctDNA; the number of mutations in each target gene is indicated. (Middle) Population prevalence of mutations in corresponding genes in breast cancer patients (METABRIC). (Right) Variant VAF (%) and predicted SnpEff functional impact. (**B**) Kaplan-Meier survival probability faceted by ctDNA detection in post-treatment samples. (**C**) Cox proportional hazards models are shown for clinical characteristics and post-treatment ctDNA status. Error bars indicate 95% confidence intervals. (**D**) Survival probability of NAT participants, faceted by ctDNA detection in post-treatment samples and whether pathologic complete response was achieved. (**E**) Intersections of pCR status, post-treatment ctDNA status and treatment with adjuvant Capecitabine in NAT participants. (**F**) Cox proportional hazards model for post-treatment ctDNA status in NAT participants who did not achieve pCR.

### Recurrence and survival probability associated with early post-treatment ctDNA

We then investigated whether patients experiencing clinical progression after treatment completion could be identified by the presence of ctDNA mutations within 7 months of completion of treatment. Cox proportional hazards analysis using clinical progression as an endpoint indicated that ctDNA positive status in the interval immediately following treatment is associated with shorter recurrence-free survival in TNBC participants (Figure 2B). Multivariable analysis showed an association between age at time of diagnosis, tumour stage, node status, and the presence of post-treatment ctDNA with participant survival and/or recurrence outcomes (post-treatment ctDNA hazard ratio (HR): 14.61, CI 2.99 - 71.27, *p* < 0.001, Figure 2C). Additionally, two covariates, specifically tumour stage T4 (HR: 15.89, CI 1.99 - 126.83, *p* = 0.009) and age at diagnosis of 50-59 years (HR: 5.243, CI 1.07 - 25.72, *p* = 0.041), were associated with participant outcomes (Figure 2C).

Since achieving pCR following NAT has been shown to be an indicator of improved long-term outcomes^35^, and 7 of the 10 participants with post-treatment ctDNA in our study received NAT, we investigated whether ctDNA status could be used as an additional prognostic indicator for participant outcomes. Progression-free survival (PFS) in participants who achieved pCR was not affected by ctDNA status (Figure 2D). For participants who did not achieve pCR following NAT, PFS was worse when ctDNA could be detected post-treatment (HR: 7.70, CI 1.80 - 32.91, *p* = 0.006), with post-treatment ctDNA detected in 4 of 40 participants with no pCR (Figure 2D-F). Additionally, 13 participants received additional Capecitabine following an incomplete response to NAT, of whom 2 had positive post-treatment ctDNA (Figure 1B and Figure 2E). Treatment with Capecitabine following an incomplete response was not associated with increased PFS, but these numbers are very small. ctDNA monitoring in serial plasma samples from a participant who did not achieve pCR and had detectable ctDNA (108) showed that although no ctDNA was detectable at time of diagnosis, a likely pathogenic TP53 variable was detectable following NAT and surgery, with higher VAF observed at the subsequent time point prior to metastatic recurrence diagnosis (Figure S2A). A similar pattern was observed for another NAT participant (109) without pCR, identified as post-treatment ctDNA positive in this study (Figure 1B). Although ctDNA was not detected in the first post-treatment sample in participant 109, the pathogenic TP53 variant R273H was detected 7 months after treatment completion, several months before metastatic recurrence diagnosis and detected again with higher VAF in plasma collected shortly after metastatic recurrence diagnosis (Figure S2A). These observations highlight the possible application of monitoring ctDNA signatures not only at different stages of treatment but during follow-up to estimate residual disease burden and identify patients with higher risk of recurrence.

Recurrence-free survival did not differ between NAT and ADJ participants, despite different rates of ctDNA detection in post-treatment samples (Figure S2B). The difference in ctDNA detection was not a marker of treatment efficacy, as both NAT and ADJ participants had overall similar survival (Table S2). Detection of post-treatment ctDNA in the ADJ group was not associated with decreased PFS (Figure S2C). However, given the low rates of ctDNA detection, significance of any associations is suggestive of a relationship and should not be used to infer patient prognosis. Four participants, 3 NAT and 1 ADJ, in our cohort had disease progression within 6 months of treatment (63-134 days, with a median of 102 days after treatment). Post-treatment ctDNA was detected in 1 NAT participant with rapidly-recurring cancer, and in 9 other participants, 3 of which had subsequent disease progression (Table S2). This observation suggests that detection of ctDNA following primary treatment completion may be informative for both rapidly-recurring disease as well as TNBC disease with clinical progression occurring more than 6 months after treatment completion and invites further investigation.

## Discussion

Pathological complete response and ctDNA status both have been demonstrated to predict survival after treatment^15,36^. While achieving pCR remains the gold standard for informing disease eradication and predicting long-term survival after NAT, lack of ctDNA detection during ongoing treatment has also been shown to better predict survival than pCR status alone^37^. ctDNA detection using patient-specific panels in plasma samples collected at different points of NAT and preceding surgery from patients with high risk early breast cancer indicated that ctDNA detection following NAT was associated with worse outcomes^16^. Conversely, ctDNA clearance at different stages of NAT was associated with recurrence-free survival, even when patients did not achieve pCR^16^. Interestingly, we observed that a combined lack of pCR and positive post-treatment ctDNA status within 7 months of treatment completion and prior to recurrence in NAT participants could be indicative of earlier TNBC disease progression due to higher residual disease burden and/or more intrinsically aggressive biology than negative pCR status alone. The ability to detect post-treatment ctDNA depends on several factors including low tumour fractions, ctDNA dynamics during treatment, and, critically, the ctDNA assay used^38^. The assay used here represents an “off the shelf” clinically validated targeted panel assay designed for a specific disease management purpose, namely the detection of actionable treatment mutations in plasma and tissue. As a pan-cancer assay targeting advanced disease, it is likely to miss a significant number of mutations of importance for TNBC monitoring as well as any patient-specific variants^38^. A recent study of exclusively residual disease burden positive patients (our study includes disease free as well as residual disease positive participants) reported a higher rate of ctDNA detection overall (33% patients) within 6 months of treatment, nevertheless emphasizing the independent prognostic value of post-treatment ctDNA^39^ with RCB scores >0 and incomplete pCR. In addition to the differences in overall study population disease burden, the higher detection rate may reflect panel size and cut-off thresholds–3 % - 40 % or > 60 % VAF compared to 1 - 40 % VAF in the current study, as well as an absence of germline sequencing for most samples^39^. In the current study, patients with relapse or clinically evident disease at completion of therapy were excluded. Moreover, we conducted extensive orthogonal validation with ddPCR and parallel sequencing of matched buffy coat samples. Recent work on ddPCR-based monitoring of moderate-high risk TNBC with a pembrolizumab intervention arm, has emphasized the need for early enrolment and sensitive detection to guide earlier intervention^40^. Including only assays negative in buffy coat and positive in tumour samples, ctDNA was detected in 27% of patients^40^. However the vast majority (>70%) had metastasis at the time of positive ctDNA detection^40^, making rates of detection more in line with those observed in our analysis of post-treatment, pre-progression plasma samples, and indicating a need for more frequent surveillance of earlier timepoints. These studies highlight the dynamic nature of ctDNA as a biomarker that can enable better prognostication. More sensitive approaches including the analysis of a large number of mutations from whole genome sequencing of plasma, in combination with the use of larger plasma volumes, or even multiparameter assays may have greater potential for implementation in future studies.

The methodology of sequencing ctDNA is an ongoing field of research, and the clinical utility of future ctDNA assays must balance genome coverage, cost, scalability, assay sensitivity and specificity and the clinical stage at which the assay is used. For the purpose of this study, a UID labelled, targeted amplicon sequencing method was used to detect a broad set of actionable SNVs in cancer. For cancers such as TNBC, where no particular “disease defining” SNVs exists other than non-specific TP53 mutations, a capture based or low-pass whole genome sequencing approach may be the better approach for ctDNA detection as they provide data on copy number changes and large structural rearrangements^41^.

ctDNA is being used successfully in a number of tumour types to assess recurrence and attempt to identify those persons who may benefit from novel targeted agents or other interventions. In our study we showed that participants who did not achieve a pCR after neoadjuvant therapy and who had detectable ctDNA had a rapid recurrence and very poor outcome. Future work using whole genome profiling methods^42,43^ across multiple timepoints following primary treatment completion will be used to identify patients with high risk of relapse and who should be enrolled in trials of early intervention with individualised therapies. Additionally, there is a need for systematic lead-time analysis between detection of post-treatment ctDNA and recurrence diagnosis. Taken together, these observations suggest that a combination of sensitive plasma based residual disease detection method in conjunction with pCR has the potential to identify, among incomplete pCR TNBC patients, those at higher risk of progression who should be evaluated for management with advanced second line therapies.

## Data Availability

Data from targeted panel sequencing are available in EGA under EGAS00001006937. Detected somatic mutations are reported in Table S3.

## Data availability

Raw data from targeted panel sequencing are available in EGA under EGAS00001006937. Detected somatic mutations are reported in Table S3.

## Author contributions

K.G. and S.A. conceptualized and oversaw the project. K.G., S.A. and W.dB. designed the project. E.Z., D.L., R.M. and R.A.-H. performed data analyses. V.C., B.Y.C.C. and A.L. contributed to sample processing, sequencing and quantification experiments. E.K., C.B., K.G., W. dB., S.B., A.C., X.L.F., D.F., A.G., N.L., C.L., S.R., T.S., C.S., S.T., and D.V. contributed to participant accrual and E.K., C.B., K.G., W. dB., and N.L. to clinical information gathering. E.K and C.B. obtained patient samples. K.T. and B.Y.C.C performed histopathology analysis. K.G., S.A., E.Z. and B.Y.C.C. wrote the manuscript. All authors contributed to manuscript editing.

## Acknowledgements

The authors gratefully acknowledge that this work would not have been possible without the participation of patients and their families. This project was supported by the BC Cancer Foundation and the Canadian Institutes of Health Research grant FDN-148429. S.A. holds the Nan and Lorraine Robertson Chair in Breast Cancer and is a Canada Research Chair in Molecular Oncology (950–230610, CRC-2021-00205). Additional funding was provided by the Susan G. Komen Leadership Grant SAB180007, and the Carolyn Baker Triple Negative Breast Cancer Fund (0BRRG004). We gratefully acknowledge the following individuals for their contributions in participant accrual: Dr. Thao Nguyen, Dr. Asif Shaikh, Dr. Wen Wen Shan, Dr. Dorothy Uhlman, and Dr. Tamana Walia from BC Cancer - Abbotsford; Dr. Simon Baxter, Dr. Kaethe Clark, Dr. Barbara Czerkawski, Dr. Susan Ellard, Dr. Suzana Gilmour, Dr. Edward Hardy, Dr. Marianne Taylor, and Dr. Delilah Topic from BC Cancer - Kelowna; Dr. Dante Wan from BC Cancer - Prince George; Dr. Theresa Chan, Dr. Balvindar Johal, Dr. Aalok Kumar, Dr. Mita Manna, Dr. Lee Ann Martin, Dr. Ravinder Sawhney, Dr. Francis Wong, and Dr. Katharine Xing from BC Cancer - Surrey; Dr. Stephen Chia, Dr. Daniel Khalaf, Dr. Sarah Lamarche, Dr. Gary Pansegrau, Dr. Moira Rushton, Dr. Amirrtha Srikanthan, Dr. Sophie Sun, and Dr. Alison Weppler from BC Cancer - Vancouver; Dr. Vanessa Bernstein, Dr. Leathia Fiorino, Dr. Doran Ksienski, Dr. Nicol Macpherson, Dr. Zia Poonja, Dr. Jeffrey Sulpher, and Dr. Joanna Vergidis from BC Cancer - Victoria.

