## Supplemental Information for "Detection of circulating plasma tumour mutations in early stage triple negative breast cancer as an adjunct to pathological complete response assessment"

**Supplementary Information**

**Table S1 -** Probes used for mutation validation using ddPCR. Probes were purchased from BioRad and the custom oligos were ordered through IDT using sequences available at BioRad for each specific probe.

| Target gene | Variant | Assay ID | Probe Fluorophore | Positive control |
|---|---|---|---|---|
| **TP53** | **R213*** | dHSACP2506810 | FAM (Mut) | Custom oligo |
| | | dHSACP2506811 | HEX (WT) | |
| | **A161T** | dHSACP2506786 | FAM (Mut) | |
| | | dHSACP2506787 | HEX (WT) | |
| | **G266E** | dHSACP2506854 | FAM (Mut) | |
| | | dHSACP2506855 | HEX (WT) | |
| | **R273C** | dHSACP2500538 | FAM (Mut) | |
| | | dHSACP2500539 | HEX (WT) | |
| | **R175H** | dHSACP200105 | FAM (Mut) | |
| | | dHSACP200106 | HEX (WT) | |
| | **C238Y** | dHSACP2506850 | FAM (Mut) | |
| | | dHSACP2506851 | HEX (WT) | |
| | **I195T** | dHSACP2506898 | FAM (Mut) | |
| | | dHSACP2506899 | HEX (WT) | |
| | **R306*** | dHSACP2500552 | FAM (Mut) | |
| | | dHSACP2500553 | HEX (WT) | |
| | **R273H** | dHSACP200109 | FAM (Mut) | |
| | | dHSACP200110 | HEX (WT) | |
| **KRAS** | **G12S** | dHSACP2500588 | FAM (Mut) | Custom oligo or Horizon HD780 multiplex ctDNA reference standard |
| | | dHSACP2500589 | HEX (WT) | |
| | **G12A** | dHSACP2500586 | FAM (Mut) | |
| | | dHSACP2500587 | HEX (WT) | |
| | **G12V** | dHSACP2500592 | FAM (Mut) | |
| | | dHSACP2500593 | HEX (WT) | |
| | **G12D** | dHSACP2500596 | FAM (Mut) | |
| | | dHSACP2500597 | HEX (WT) | |
| | **G12C** | dHSACP2500584 | FAM (Mut) | Custom oligo |
| | | dHSACP2500585 | HEX (WT) | |

| | | | | |
|---|---|---|---|---|
| **PIK3CA** | **E545K** | dHSACP2000075 | FAM (Mut) | MCF-7 cell line or Horizon HD780 multiplex ctDNA reference standard |
| | | dHSACP2000076 | HEX (WT) | |
| | **E542K** | dHSACP2000073 | FAM (Mut) | Custom oligo |
| | | dHSACP2000074 | HEX (WT) | |
| | **H1047R** | dHSACP2000077 | FAM (Mut) | HCT116 cell line |
| | | dHSACP2000078 | HEX (WT) | |
| **IDH1** | **R132H** | dHSACP200055 | FAM (Mut) | Custom oligo |
| | | dHSACP200056 | HEX (WT) | |
| | **R132G** | dHsaMDV2510512 | FAM/HEX (Mut/WT) | Custom oligo |
| **NRAS** | **Q61R** | dHSACP200071 | FAM (Mut) | Custom oligo |
| | | dHSACP200072 | HEX (WT) | |
| **BRAF** | **V600E** | dHSACP200027 | FAM (Mut) | Custom oligo |
| | | dHSACP200028 | HEX (WT) | |
| **POLE** | **T457T** | dHsaMDS230604489 | FAM/HEX (Mut/WT) | Custom oligo |
| | **T457M** | dHsaMDS195642452 | FAM/HEX (Mut/WT) | Custom oligo |
| **EGFR** | **A722V** | dHsaMDS839693386 | FAM/HEX (Mut/WT) | Custom oligo |

**Table S2**. Clinical characteristics of participants in cohort, and their comparison between NAT and ADJ participants, indicated by p-values for chi-square (categorical variables) and Wilcoxon tests (continuous variables). A *p*-value < 0.05 was considered statistically significant.

| | | **All (n=130)** | **NAT (n=64)** | **Adjuvant (n=66)** | ***p*-value** |
|---|---|---|---|---|---|
| Age, in years, at diagnosis range (median) | | 24 - 78 (54) | 24 - 76 (52) | 32 - 78 (56) | 0.08 |
| Node status (%) | Positive | 53 (40.8) | 35 (54.7) | 18 (27.3) | 0.0009 |
| | Negative | 75 (57.7) | 27 (42.2) | 48 (72.7) | |
| | Unknown | 2 (1.5) | 2 (3.1) | - | |
| LVI (%) | Present | 20 (15.4) | 6 (9.4) | 14 (21.2) | 0.01 |
| | Not identified | 97 (74.6) | 48 (75.0) | 49 (74.2) | |
| | Suspicious | 3 (2.3) | 1 (1.6) | 2 (3.0) | |
| | Indeterminate | 1 (0.8) | - | 1 (1.5) | |
| | Unknown | 9 (6.9) | 9 (14.1) | - | |
| Stage (%) | T1 | 38 (29.2) | 10 (15.6) | 28 (42.4) | 8e-05 |
| | T2 | 65 (50.0) | 32 (50.0) | 33 (50.0) | |
| | T3 | 22 (16.9) | 17 (26.6) | 5 (7.6) | |
| | T4 | 5 (3.9) | 5 (7.8) | - | |
| Grade (%) | 2 | 27 (20.8) | 14 (21.9) | 13 (19.7) | 0.9 |
| | 3 | 102 (78.5) | 50 (78.1) | 52 (78.8) | |
| | Unknown | 1 (0.8) | - | 1 (1.5) | |
| Tumour size, in cm, range (median) | | 0.25 - 9.30 (2.6) | 0.25 - 8.00 (3.4) | 0.70 - 9.30 (2.2) | 9e-06 |
| Clinical genetics (%) | Negative | 56 (43.1) | 29 (45.3) | 27 (40.9) | 0.9 |
| | Variant identified | 27 (20.8) | 14 (21.9) | 13 (19.7) | |
| | VUS | 20 (15.4) | 9 (14.1) | 11 (16.7) | |
| | Not done | 27 (20.8) | 12 (18.8) | 15 (22.7) | |
| Median interval between treatment end and post-treatment sample (months) | | 1.8 | 1.7 | 2.4 | 0.6 |
| Recurrence or death (%) | | 17 (13.1) | 10 (15.6) | 7 (10.6) | 0.4 |
| Median interval between treatment end and recurrence or death (months) | | 10.8 | 6.8 | 16.3 | 0.2 |
| Treatment type (%) | Triple therapy, anthracycline | 68 (52.3) | 26 (40.6) | 42 (63.6) | 1e-09 |

| | | | | | |
|---|---|---|---|---|---|
| | and alkylating, then taxane | | | | |
| | Dual therapy | 20 (15.4) | 2 (3.1) | 18 (27.3) | |
| | Triple therapy, taxane first, then anthracycline and alkylating | 31 (23.8) | 27 (42.2) | 4 (6.1) | |
| | Protocol contains platinum | 10 (7.7) | 9 (14.1) | 1 (1.5) | |
| | Other | 1 (0.8) | - | 1 (1.5) | |
| Radiation (%) | Yes | 113 (86.9) | 60 (93.8) | 53 (80.3) | 0.05 |
| | No | 16 (12.3) | 4 (6.2) | 12 (18.1) | |
| | Other | 1 (0.8) | - | 1 (1.5) | |
| Pathologic complete response in NAT participants (%) | Yes | - | 24 (37.5) | - | - |
| | No | - | 40 (62.5) | - | |

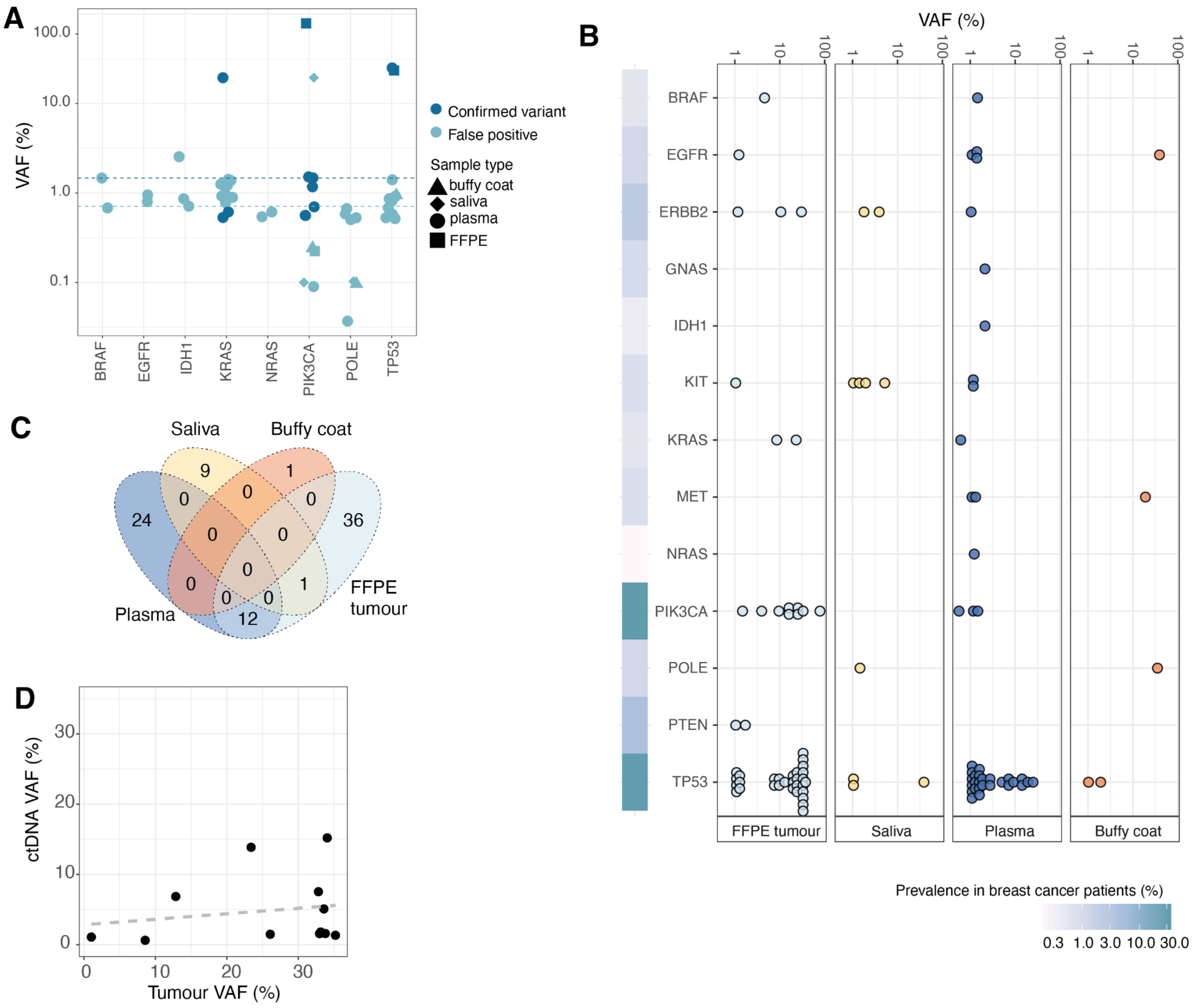


**Figure S1**.(**A**) Distribution of VAF (%) across genes for variants validated by digital droplet PCR (ddPCR). The dashed lines represent the median VAF (%) for confirmed variants (dark blue) and false positive variants (light blue). (**B**) Comparison of VAF (%) in genes containing detectable mutations by sample type; breast cancer patient population (METABRIC and TCGA) prevalence of mutations in these genes are indicated by the heatmap. (**C**) Participant-specific overlap of variants in different sample types to identify likely germline variants and evaluate clonal hematopoiesis. (**D**) Correlation of VAFs for variants identified in participant-matched tumour and plasma samples.

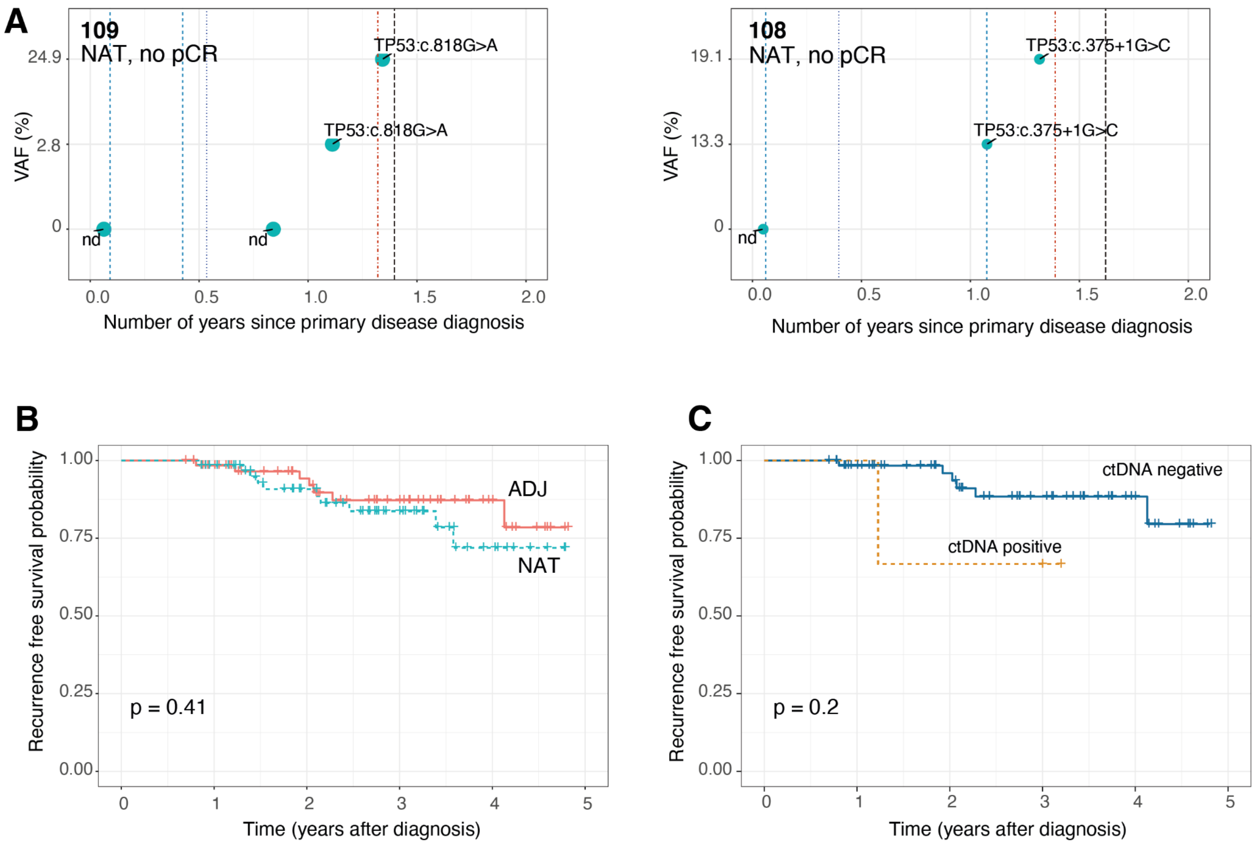


**Figure S2.** (**A**) Variants detected in serial plasma samples from 2 NAT participants with an incomplete response. Circles represent targeted panel-sequenced plasma samples, red dashed line represents recurrence diagnosis, blue dashed lines are the first and last chemo treatments, and the purple dotted line indicates date of surgical resection. (**B**) Recurrence-free survival in participants in cohort, faceted by NAT or ADJ chemotherapy, and (**C**) recurrence-free survival in participants receiving ADJ chemotherapy, faceted by ctDNA status.

**Table S3**. Somatic variants detected in plasma and FFPE tumour samples.

| Gene name | RefSeq ID | cDNA change | Strand | Mutation type | Predicted impact (SnpEff) |
|---|---|---|---|---|---|
| BRAF | NM_004333.4 | c.1397G>T | -1 | missense_variant | MODERATE |
| BRAF | NM_004333.4 | c.1780G>A | -1 | missense_variant | MODERATE |
| EGFR | NM_005228.3 | c.2139A>G | 1 | synonymous_variant | LOW |
| EGFR | NM_005228.3 | c.2327G>A | 1 | missense_variant | MODERATE |
| EGFR | NM_005228.3 | c.2458C>T | 1 | stop_gained | HIGH |
| EGFR | NM_005228.3 | c.2611G>A | 1 | missense_variant | MODERATE |
| ERBB2 | NM_004448.3 | c.2329G>T | 1 | missense_variant | MODERATE |
| ERBB2 | NM_004448.3 | c.2463C>T | 1 | synonymous_variant | LOW |

| | | | | | |
|---|---|---|---|---|---|
| ERBB2 | NM_004448.3 | c.2464C>T | 1 | synonymous_variant | LOW |
| ERBB2 | NM_004448.3 | c.929C>T | 1 | missense_variant | MODERATE |
| GNAS | NM_080425.3 | c.2531G>A | 1 | missense_variant | MODERATE |
| IDH1 | NM_001282386.1 | c.394C>G | -1 | missense_variant | MODERATE |
| KIT | NM_000222.2 | c.1497G>A | 1 | synonymous_variant | LOW |
| KIT | NM_000222.2 | c.1753C>T | 1 | missense_variant | MODERATE |
| KRAS | NM_033360.3 | c.35G>A | -1 | missense_variant | MODERATE |
| KRAS | NM_033360.3 | c.35G>T | -1 | missense_variant | MODERATE |
| MET | NM_001127500.2 | c.2847C>T | 1 | synonymous_variant | LOW |
| MET | NM_001127500.2 | c.2883A>G | 1 | synonymous_variant | LOW |
| NRAS | NM_002524.4 | c.182A>G | -1 | missense_variant | MODERATE |
| PIK3CA | NM_006218.3 | c.1624G>A | 1 | missense_variant | MODERATE |
| PIK3CA | NM_006218.3 | c.1633G>A | 1 | missense_variant | MODERATE |
| PIK3CA | NM_006218.3 | c.3140A>G | 1 | missense_variant | MODERATE |
| PTEN | NM_001304717.2 | c.908G>A | 1 | missense_variant | MODERATE |
| TP53 | NM_000546.5 | c.105G>A | -1 | synonymous_variant | LOW |
| TP53 | NM_000546.5 | c.167A>G | -1 | missense_variant | MODERATE |
| TP53 | NM_000546.5 | c.181G>A | -1 | missense_variant | MODERATE |
| TP53 | NM_000546.5 | c.212C>A | -1 | missense_variant | MODERATE |
| TP53 | NM_000546.5 | c.242C>T | -1 | missense_variant | MODERATE |
| TP53 | NM_000546.5 | c.309C>T | -1 | synonymous_variant | LOW |
| TP53 | NM_000546.5 | c.331C>T | -1 | synonymous_variant | LOW |
| TP53 | NM_000546.5 | c.332T>C | -1 | missense_variant | MODERATE |
| TP53 | NM_000546.5 | c.374C>T | -1 | missense_variant&splice_region_variant | MODERATE |
| TP53 | NM_000546.5 | c.375+1G>C | -1 | splice_donor_variant&intron_variant | HIGH |
| TP53 | NM_000546.5 | c.378C>T | -1 | splice_region_variant&synonymous_variant | LOW |
| TP53 | NM_000546.5 | c.413C>T | -1 | missense_variant | MODERATE |
| TP53 | NM_000546.5 | c.476C>T | -1 | missense_variant | MODERATE |
| TP53 | NM_000546.5 | c.488A>G | -1 | missense_variant | MODERATE |
| TP53 | NM_000546.5 | c.527G>A | -1 | missense_variant | MODERATE |

| | | | | | |
|---|---|---|---|---|---|
| TP53 | NM_000546.5 | c.584T>C | -1 | missense_variant | MODERATE |
| TP53 | NM_000546.5 | c.587G>A | -1 | missense_variant | MODERATE |
| TP53 | NM_000546.5 | c.604C>T | -1 | missense_variant | MODERATE |
| TP53 | NM_000546.5 | c.614A>G | -1 | missense_variant | MODERATE |
| TP53 | NM_000546.5 | c.637C>T | -1 | stop_gained | HIGH |
| TP53 | NM_000546.5 | c.639A>G | -1 | synonymous_variant | LOW |
| TP53 | NM_000546.5 | c.641A>G | -1 | missense_variant | MODERATE |
| TP53 | NM_000546.5 | c.646G>A | -1 | missense_variant | MODERATE |
| TP53 | NM_000546.5 | c.713G>A | -1 | missense_variant | MODERATE |
| TP53 | NM_000546.5 | c.722C>T | -1 | missense_variant | MODERATE |
| TP53 | NM_000546.5 | c.743G>A | -1 | missense_variant | MODERATE |
| TP53 | NM_000546.5 | c.782+1G>A | -1 | splice_donor_variant& intron_variant | HIGH |
| TP53 | NM_000546.5 | c.782+2T>G | -1 | splice_donor_variant& intron_variant | HIGH |
| TP53 | NM_000546.5 | c.801G>C | -1 | synonymous_variant | LOW |
| TP53 | NM_000546.5 | c.813G>A | -1 | synonymous_variant | LOW |
| TP53 | NM_000546.5 | c.816G>C | -1 | synonymous_variant | LOW |
| TP53 | NM_000546.5 | c.817C>T | -1 | missense_variant | MODERATE |
| TP53 | NM_000546.5 | c.818G>A | -1 | missense_variant | MODERATE |
| TP53 | NM_000546.5 | c.818G>T | -1 | missense_variant | MODERATE |
| TP53 | NM_000546.5 | c.819T>A | -1 | synonymous_variant | LOW |
| TP53 | NM_000546.5 | c.825T>C | -1 | synonymous_variant | LOW |
| TP53 | NM_000546.5 | c.832C>A | -1 | missense_variant | MODERATE |
| TP53 | NM_000546.5 | c.839G>A | -1 | missense_variant | MODERATE |
| TP53 | NM_000546.5 | c.841G>C | -1 | missense_variant | MODERATE |
| TP53 | NM_000546.5 | c.843C>A | -1 | missense_variant | MODERATE |
